# Azithromycin for infants at risk of poor growth and development: a pooled secondary analysis of two randomized controlled trials

**DOI:** 10.1101/2025.04.27.25326525

**Authors:** Mamadou Bountogo, Mamadou Ouattara, Clarisse Dah, Boubacar Coulibaly, Thierry Ouedraogo, Alphonse Zakane, Valentin Boudo, Elodie Lebas, Huiyu Hu, Benjamin F. Arnold, Thomas M. Lietman, Ali Sié, Catherine E. Oldenburg

## Abstract

**Background:** In 2023, the World Health Organization (WHO) revised its guidelines for management of severe acute malnutrition (SAM). The revised guidelines include a focus on infants at risk of poor growth and development. The guideline identifies evaluation of routine antibiotics for these infants as a priority research area.

**Objective:** We pooled data from two large randomized controlled trials evaluating azithromycin for prevention of infant mortality in Burkina Faso to assess whether azithromycin reduces mortality or wasting in this subgroup.

**Methods:** Infants in the two trials were 1-12 weeks of age at enrollment. Infants were considered at risk of poor risk of growth and development per WHO: underweight (weight-for-age Z-score, WAZ < −2), wasted (weight-for-length Z-score, WLZ < −2), or MUAC < 11.0 cm among infants ≥6 weeks of age. Infants were randomized to a single oral (20 mg/kg) dose of azithromycin or matching placebo and were followed until 6 months of age. We evaluated vital status, underweight (WAZ < −2), wasting (WLZ < −2), and stunting (length-for-age Z-score, LAZ) at 6 months among infants at risk of poor growth and development based on WHO single measurement criteria.

**Results:** A total of 54,709 infants were enrolled in the two trials. Of these, 9,728 were at risk of poor growth and development based on baseline WAZ (N=5,385), WLZ (N=6,022), or MUAC (N=1,541). We found no evidence of a difference in mortality (1.3% vs 1.1%, odds ratio, OR, 1.19, 95% confidence interval, CI, 0.82 to 1.72) or wasting (20.6% vs 20.2%, OR 1.03, 95% CI 0.92 to 1.14) at 6 months among infants receiving azithromycin versus placebo.

**Conclusions:** In infants aged 1-12 weeks at risk of poor growth and development, we do not have evidence that single dose azithromycin reduces mortality or improves growth outcomes.

**Trial Registration:** ClinicalTrials.gov NCT03682654 and NCT03676764

## INTRODUCTION

The majority of the evidence base for the management of malnutrition in children is focused on children aged 6 to 59 months. Considerably less evidence exists to support treatment severe wasting in infants < 6 months old, and current treatment guidelines include inpatient care for infants with severe acute malnutrition.^1^ Recent evidence has demonstrated that the highest incidence of wasting may be during early infancy^2^, and thus evidence-based interventions for treatment and prevention in infants under 6 months of age may be the most impactful for reducing the harmful effects of wasting in children.

In 2023, the World Health Organization (WHO) revised its guidelines for the prevention and management of wasting and included increased emphasis on wasting in infants < 6 months of age.^3^ In the revised guidelines, WHO identified a new group of infants, infants at risk of poor growth and development, who are a priority for identification and intervention to prevent progression to wasting. WHO defines this group in several ways, including poor growth based on sequential measures as well as with poor anthropometric measurements based on a single measure. WHO identified a number of priority research questions for the management of infants at risk of poor growth and development, including whether antibiotics should be routinely administered to infants at risk of poor growth and development.

Provision of routine antibiotics is included in WHO guidelines as part of the management of uncomplicated severe wasting in children aged 6 to 59 months.^3^ Antibiotics may improve outcomes for children with severe wasting as these children may have compromised immune systems and subclinical infections that adversely affect their probability of recovery. Most treatment guidelines include a course of amoxicillin or other broad-spectrum antibiotic for children treated on an outpatient basis. However, the evidence base for inclusion of amoxicillin in treatment guidelines is mixed.^4,5^ A trial in Malawi found that children receiving amoxicillin or ceftriaxone had significantly reduced mortality and improved recovery^5^, however a trial in Niger found no impact of routine amoxicillin on probability of recovery.^4^

Mass distribution of single dose azithromycin has been shown to reduce all-cause child mortality among children 1-59 months.^6,7^ However, single dose azithromycin delivered to infants aged 1 to 12 weeks regardless of nutritional status has not been shown to reduce mortality or improve growth outcomes by 6 months of age in two trials in Burkina Faso.^8–11^ In a subgroup analysis of the neonatal azithromycin trial, we did not find evidence to support an effect of azithromycin in most subgroups of neonates defined by low anthropometric measures (e.g., wasting, stunting), but infants who were both low birthweight and underweight at the time of enrollment (up to 28 days of age) who received azithromycin had reduced odds of underweight at 6 months of age compared to placebo.^12^ Azithromycin has some advantage over other antibiotic classes, as it can be dosed as a single oral dose due to its long half-life. Here, we pooled data from the NAITRE and CHATON trials^8,9^, two placebo-controlled trials of azithromycin for prevention of mortality in early infancy that followed nearly identical protocols, to provide evidence of the role of azithromycin for prevention of mortality, wasting, underweight, and stunting in infants at risk of poor growth and development at enrollment.

## METHODS

### Study setting

Both NAITRE and CHATON were conducted in the Boucle du Mouhoun, Hauts-Bassins, and Cascades regions of Burkina Faso. Participants in NAITRE were additionally enrolled in Centre and Centre Ouest regions. Burkina Faso is a landlocked country in the Sahel region of West Africa that experiences highly seasonal rainfall, from approximately July through October. Participants were enrolled at *Centres de Santé et de Promotion Sociale* (CSPS), which are government-run primary healthcare facilities that deliver first-line preventative and curative care. Healthcare is free for children under five years of age at these facilities.

### Parent Trial Methods

Complete methods for the parent trials have been previously reported.^7,9,13,14^ In brief, the NAITRE and CHATON trials were 1:1 randomized placebo-controlled trials designed to evaluate the efficacy of a single oral 20 mg/kg dose of azithromycin compared to matching placebo for prevention of all-cause mortality. Neither trial demonstrated an effect of azithromycin compared to placebo for mortality.^7,9^ In both trials, participants were followed until 6 months of age. The trials were reviewed and approved by the Institutional Review Board at the University of California, San Francisco, the Comité National d’Ethique pour la Recherche, and the Comité Technique d’Examen des Demandes d’Autorisation d’Essais Clinique in Ouagadougou, Burkina Faso. Written informed consent was obtained from at least one guardian of each enrolled infant. Both trials were registered at ClinicalTrials.gov (NAITRE: NCT03682654; CHATON: NCT03676764).

### Participants

In NAITRE, infants were eligible for the trial if they were between 8 and 27 days of age, weighed at least 2500 g at the time of enrollment, were able to feed orally, had no known allergies to macrolides, and had no evidence of neonatal jaundice at enrollment based on clinical signs. Infants < 2500 g were excluded from NAITRE due to hypothesized increased risk of infantile hypertrophic pyloric stenosis among underweight infants.^15^ Infants were recruited via recruiting women who gave birth at participating facilities and vaccination visits during the first month. Infants in CHATON were eligible if they were between 5 and 12 weeks of age, were able to orally feed, and had no known allergies to macrolides. There were no weight-based exclusion criteria in CHATON because infants were older at the time of enrollment. Infants were recruited via well-infant visits in the clinic as well as via community-based outreach for vaccination. Participants in NAITRE were enrolled from April 2019 through December 2020 and participants in CHATON were enrolled from September 2019 through December 2022.

### Intervention, randomization, and masking

Infants were randomly assigned in a 1:1 fashion with no stratification or blocking. Infants randomized to azithromycin received a single directly observed dose of 20 mg/kg azithromycin. Those randomized to placebo received an equivalent volume of matching placebo. Participants, caregivers, trial staff, and investigators were masked to the infant’s randomized treatment assignment.

### Vital status assessment

The primary outcome for both trials was all-cause mortality by 6 months of age. Vital status was assessed via caregiver visit at the clinic at 6 months of age. If a caregiver did not return, attempts were made to conduct a home visit or assess vital status via phone call.

### Anthropometric measures

In both trials, anthropometric measures were collected at enrollment and at the 6-month visit.^10,11^ Equipment and protocols for anthropometric measures were the same in both studies. At both study visits, weight was measured using a digital infant scale (ADE M112600U scale, Hamburg, Germany). Length was measured using a ShorrBoard (Weigh and Measure, Olney, MD). Mid-upper arm circumference (MUAC) measurements were collected using standard MUAC tapes. WLZ, weight-for-age Z-score (WAZ), and length-for-age Z-score (LAZ) were calculated using 2006 WHO reference standards.^16^ Infants were classified as “at risk of poor growth and development” according to WHO criteria for single measures at the time of enrollment: 1) WLZ < −2, 2) WAZ < - 2, or 3) MUAC < 11.0 for infants ≥ 6 weeks of age. At 6 months, wasting, underweight, and stunting were defined as WLZ < −2, WAZ < −2, and LAZ < −2, respectively.

### Statistical methods

To assess the risk of mortality, wasting, underweight, and stunting at 6 months among infants at risk of poor growth and development compared to those not classified as at risk of poor growth and development, we used logistic regression models with a term for risk category (e.g., “at risk” or not) adjusting for the child’s age in days at enrollment, sex, region, and season of enrollment for each outcome. A separate logistic regression model for each individual indicator (e.g., low WLZ, low WAZ, and low MUAC) was then run to assess associations between each indicator and mortality, wasting, underweight, and stunting at 6 months. For the low MUAC indicator, the analysis was restricted only to infants ≥ 6 weeks of age, as there are no established MUAC-based criteria for classifying infants as being at risk of poor growth and development based on MUAC who are <6 weeks of age.

An intention-to-treat approach was taken for all randomized comparisons. To evaluate the effect of azithromycin on mortality, wasting, underweight, and stunting in infants at risk of poor growth and development, we used logistic regression models with a term for the child’s randomized treatment assignment with a fixed effect for the parent trial. For continuous measures of WLZ, WAZ, and LAZ at 6 months, a linear regression model was used for each outcome, with a term for the infant’s randomized treatment assignment and a fixed effect for the parent study. A similar analytic strategy was taken to evaluate the effect of azithromycin in each individual indicator of risk (low WLZ, low WAZ, and low MUAC), with low MUAC models restricted only to infants ≥ 6 weeks of age as previously described. All analyses were conducted in Stata 17.0 (StataCorp, College Station, TX). All analyses were exploratory and were not pre-specified in the original trials’ statistical analysis plans.

## RESULTS

Of 54,709 infants enrolled in the two trials, 9,728 (18%) were classified as at risk of poor growth and development at baseline based on WLZ, WAZ, and/or MUAC. Of these, 4,913 were in the azithromycin group and 4,815 in the placebo group (**Figure 1**). In the azithromycin group, 4,792 (98%) and 4,242 (86%) were including in mortality and anthropometric analyses at 6 months. In the placebo group, 4,627 (96%) and 4,201 (87%) were included in mortality and anthropometric analyses at 6 months.

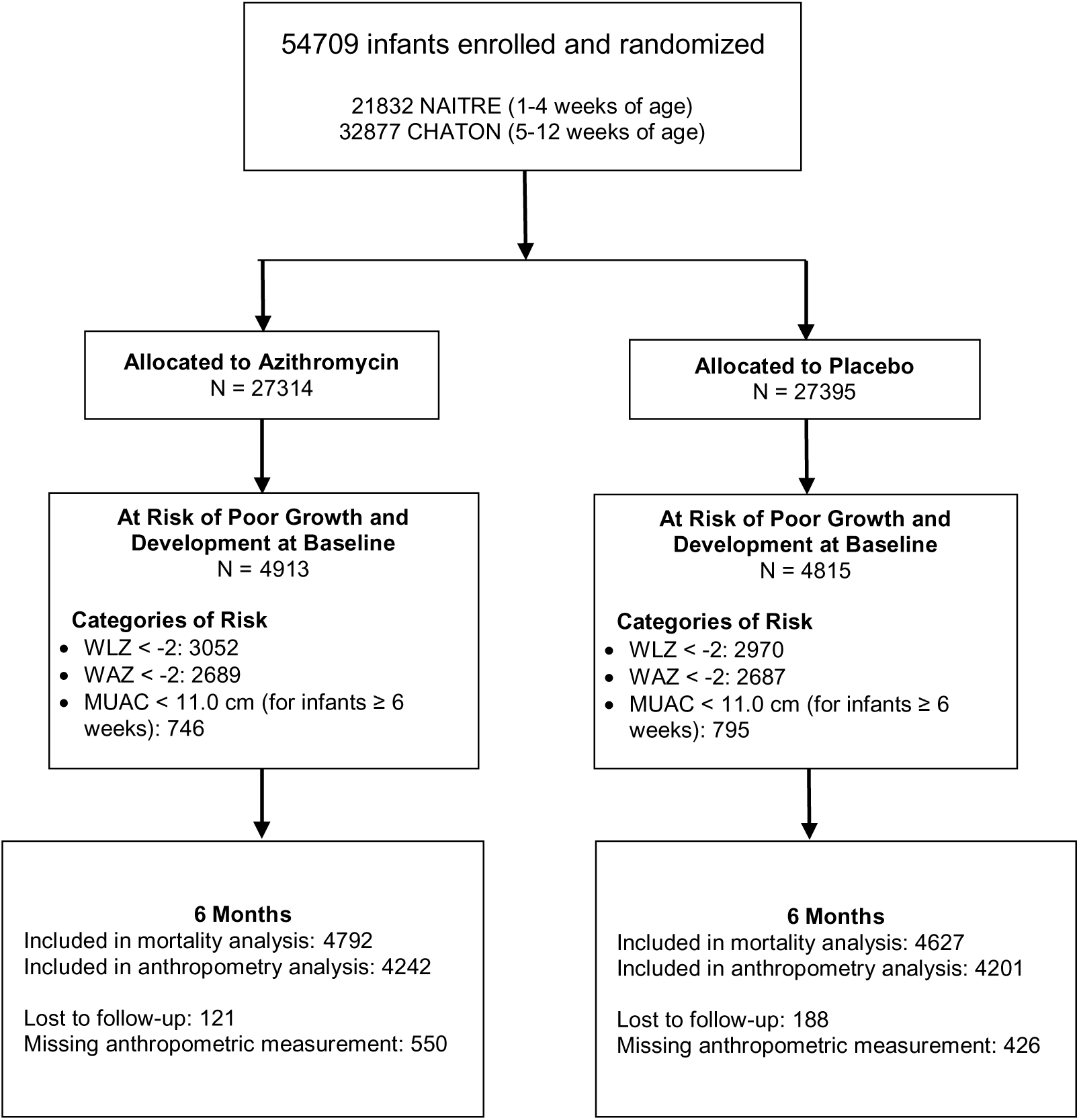

Among infants at risk of poor growth and development, baseline characteristics were well balanced between treatment groups overall and within cohorts (**Table 1**). Overall, slightly more than half of participants were male and median age was 33 days at baseline. WLZ was the most common indicator for identifying infants at risk of poor growth and development overall and was more common among younger infants enrolled in NAITRE compared to older infants in CHATON.

**Table 1.**
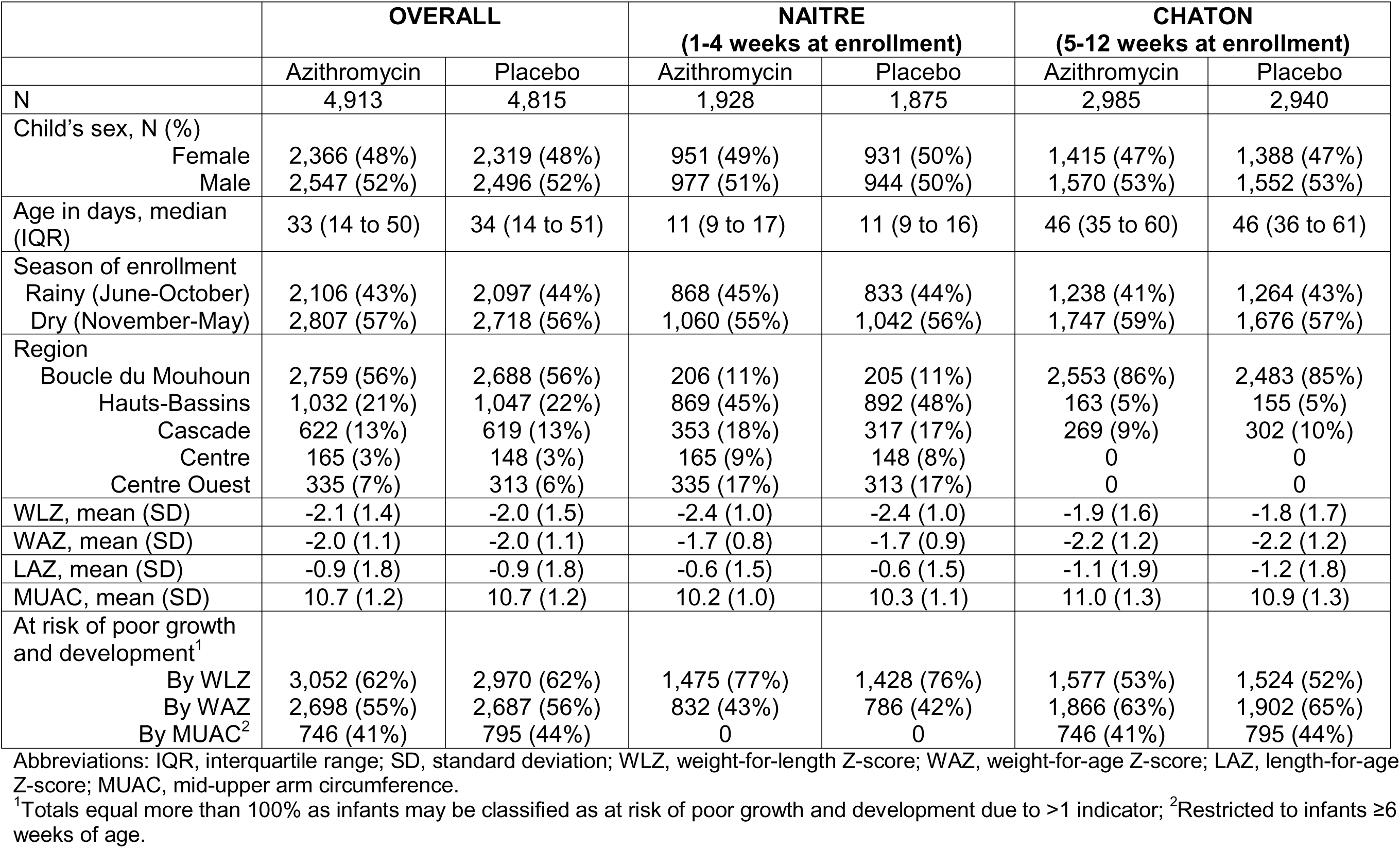
Baseline characteristics of infants at risk of poor growth and development as defined by weight-for-length Z-score, weight-for-age Z-score, and mid-upper arm circumference by randomized treatment assignment in the NAITRE (age 1-4 weeks) and CHATON (age 5-12 weeks) cohorts.

By 6 months of age, 1.2% of infants classified at risk of poor growth and development had died, compared to 0.3% not classified as at risk of poor growth and development (odds ratio, OR, for mortality 3.78, 95% confidence interval, CI, 3.06 to 4.66; **Table 2**). Wasting prevalence at 6 months was 20.4% in infants at risk of poor growth and development compared to 7.8% in those not at risk of poor growth and development (OR for wasting 3.05, 95% CI 2.21 to 4.19; **Table 2**). The odds of underweight and stunting were higher in infants at risk of poor growth and development compared to those not classified as at risk of poor growth and development (**Table 2**). Results were similar for individuals with each individual indicator of risk of poor growth and development compared to those without the indicator (e.g., low WLZ, low WAZ, and low MUAC, **Table 2**). Infants at risk of poor growth and development as classified by WAZ < −2 and MUAC < 11.0 had the highest risk of mortality (1.7% and 2.1%, respectively) and wasting (22.2% and 27.5%, respectively) at 6 months (**Figure 2**).

**Table 2.** Risk of mortality, wasting, underweight, and stunting at 6 months of age among infants at risk of poor growth in development compared to those not at risk of poor growth and development^1^.

|  | **At risk<sup>1</sup>** |  | **Not at risk** |  | **OR<sup>5</sup> (95% CI)** |
| --- | --- | --- | --- | --- | --- |
|  | *n with outcome (%)* | *N in analysis* | *n with outcome (%)* | *N in analysis* |  |
| Died | 115 (1.2%) | 9,471 | 142 (0.3%) | 43,635 | 3.78 (3.06 to 4.66) |
| Wasted<sup>2</sup> | 1,721 (20.4%) | 8,443 | 3,046 (7.8%) | 38,946 | 3.05 (2.21 to 4.19) |
| Underweight<sup>3</sup> | 2,104 (24.9%) | 8,443 | 2,900 (7.5%) | 38,946 | 4.21 (2.11 to 8.40) |
| Stunted<sup>4</sup> | 1,545 (18.3%) | 8,443 | 2,771 (7.1%) | 38,946 | 2.98 (1.32 to 6.70) |
|  | **At risk by WLZ** |  | **Not at risk by WLZ** |  | **OR<sup>5</sup> (95% CI)** |
| Died | 66 (1.1%) | 5,873 | 191 (0.4%) | 47,233 | 2.84 (2.55 to 3.16) |
| Wasted<sup>2</sup> | 1,149 (21.7%) | 5,296 | 3,618 (8.6%) | 42,093 | 3.19 (2.22 to 4.58) |
| Underweight<sup>3</sup> | 1,028 (19.4%) | 5,296 | 3,976 (9.5%) | 42,093 | 2.50 (1.55 to 4.01) |
| Stunted<sup>4</sup> | 583 (11.0%) | 5,296 | 3,733 (8.9%) | 42,093 | 1.32 (0.84 to 2.09) |
|  | **At risk by WAZ** |  | **Not at risk by WAZ** |  | **OR<sup>5</sup> (95% CI)** |
| Died | 89 (1.7%) | 5,252 | 168 (0.4%) | 47,854 | 4.93 (3.91 to 6.22) |
| Wasted<sup>2</sup> | 1,027 (22.2%) | 4,629 | 3,740 (8.8%) | 42,760 | 2.81 (2.09 to 3.78) |
| Underweight<sup>3</sup> | 1,642 (35.5%) | 4,629 | 3,362 (7.9%) | 42,760 | 6.17 (3.44 to 11.1) |
| Stunted<sup>4</sup> | 1,301 (28.1%) | 4,629 | 3,015 (7.1%) | 42,760 | 5.10 (2.57 to 10.1) |
|  | **At risk by MUAC<sup>6</sup>** |  | **Not at risk by MUAC<sup>6</sup>** |  | **OR<sup>7</sup> (95% CI)** |
| Died | 32 (2.1%) | 1,504 | 63 (0.4%) | 17,740 | 6.15 (3.96 to 9.54) |
| Wasted<sup>2</sup> | 358 (27.5%) | 1,303 | 1,695 (11.0%) | 15,364 | 3.39 (2.96 to 3.88) |
| Underweight<sup>3</sup> | 518 (39.8%) | 1,303 | 1,659 (10.8%) | 15,364 | 6.30 (5.55 to 7.16) |
| Stunted<sup>4</sup> | 382 (29.3%) | 1,303 | 1,201 (7.8%) | 15,364 | 5.20 (4.53 to 5.97) |

Abbreviations: OR, odds ratio; WLZ, weight-for-length Z-score; WAZ, weight-for-age Z-score; MUAC, mid-upper arm circumference
<sup>1</sup>Infants were determined to be at risk of poor growth and development per World Health Organization criteria for single timepoint measures (weight-for-length Z-score < -2 and/or weight-for-age < -2 and/or mid-upper arm circumference < 11.0 for infants > 6 weeks of age); <sup>3</sup>Weight-for-length Z-score < -2;
<sup>3</sup>Weight-for-age Z-score < -2; <sup>4</sup>Length-for-age Z-score < -2; <sup>5</sup>Logistic regression model with standard errors accounting for clustering by parent trial and adjusting for the child's age in days, sex, region, and season of enrollment; <sup>6</sup>Restricted only to children > 6 weeks of age; <sup>7</sup>Analysis restricted only to infants in CHATON ≥6 weeks of age due to age eligibility, logistic regression model adjusting for the child's age in days, sex, region, and season of enrollment.

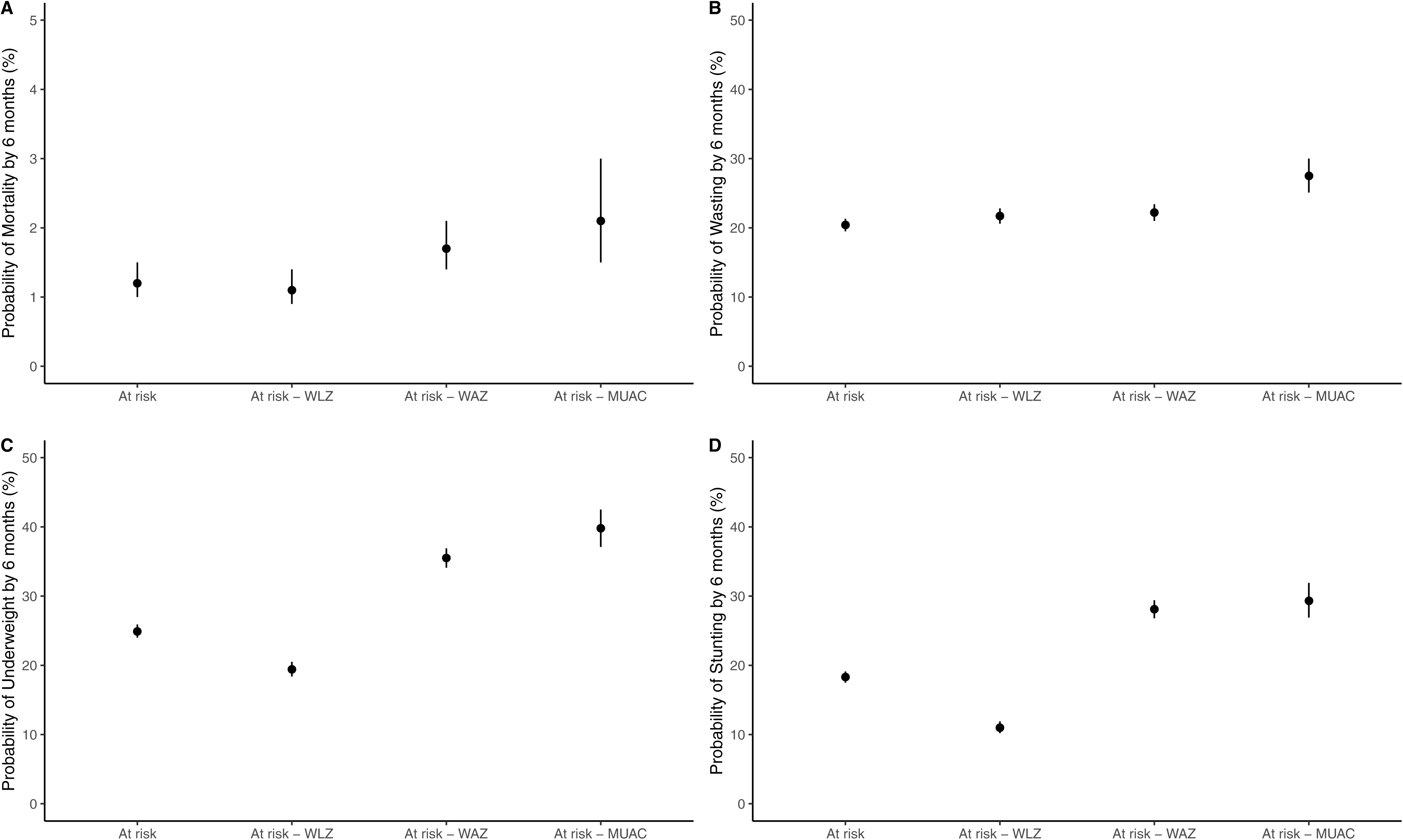

We found no evidence of an effect of azithromycin compared to placebo on mortality, wasting, underweight, or stunting at 6 months among infants at risk of poor growth and development by all indicators or by any specific indicator (**Table 3**; **Figure 3**). By 6 months of age, 1.3% of infants in the azithromycin group died compared to 1.1% in the placebo group (OR 1.19, 95% CI 0.82 to 1.72, *P*=0.36). Approximately 20% of infants were wasted at 6 months in both groups (azithromycin: 20.6%; placebo: 20.2%; OR 1.03, 95% CI 0.92 to 1.14, *P*=0.63). There was no evidence of a difference in underweight (OR 0.96, 96% CI 0.87 to 1.06, *P*=0.43) or stunting (OR 0.94, 95% CI 0.84 to 1.05, *P*=0.25) in infants receiving azithromycin compared to placebo. As continuous measures, WLZ and WAZ were similar in the azithromycin and placebo groups (WLZ: mean difference −0.03 SD, 95% CI −0.09 to 0.3, *P*=0.26; WAZ: mean difference 0.02 SD, 95% CI −0.02 to 0.07, *P*=0.54; **Table 3**). LAZ at 6 months was modestly higher in infants receiving azithromycin compared to placebo (mean difference 0.07 SD, 95% CI 0.008 to 0.13, *P*=0.03).

**Table 3.** Effect of azithromycin compared to placebo on mortality, wasting, underweight, and stunting at 6 months of age among infants classified as at risk of poor growth and development^1^ at weeks 1-12 of age.

|  | **Azithromycin**<br>**N with outcome**<br>**(%)** | **Placebo**<br>**N with outcome**<br>**(%)** | **OR<sup>7</sup> (95% CI)** | **P-value** |
| --- | --- | --- | --- | --- |
| Died<sup>2</sup> | 63 (1.3%) | 52 (1.1%) | 1.19 (0.82 to 1.72) | 0.36 |
| Wasted<sup>3,4</sup> | 872 (20.6%) | 849 (20.2%) | 1.03 (0.92 to 1.14) | 0.63 |
| Underweight<sup>3,5</sup> | 1,040 (24.5%) | 1,064 (25.3%) | 0.96 (0.87 to 1.06) | 0.43 |
| Stunted<sup>3,6</sup> | 755 (17.8%) | 790 (18.8%) | 0.94 (0.84 to 1.05) | 0.25 |
|  | **Mean (SD)** | **Mean (SD)** | **Mean Difference**<br>**(95% CI)** | **P-value** |
| WLZ | -0.93 (1.4) | -0.90 (1.5) | -0.03 (-0.09 to 0.3) | 0.26 |
| WAZ | -1.2 (1.3) | -1.2 (1.3) | 0.02 (-0.04 to 0.07) | 0.54 |
| LAZ | -0.70 (1.5) | -0.77 (1.5) | 0.07 (0.008 to 0.13) | 0.03 |

Abbreviations: OR, odds ratio; WLZ, weight-for-length Z-score; WAZ, weight-for-age Z-score; LAZ, length-for-age Z-score.
<sup>1</sup>Infants were determined to be at risk of poor growth and development per World Health Organization criteria for single timepoint measures (weight-for-length Z-score < -2 and/or weight-for-age < -2 and/or mid-upper arm circumference < 11.0 for infants > 6 weeks of age; <sup>2</sup>N=9,471 in analysis; <sup>3</sup>N=8,443 in analysis; <sup>4</sup>Defined as weight-for-length Z-score < -2; <sup>5</sup>defined as weight-for-age Z-score < -2; <sup>6</sup>defined as length-for-age Z-score < -2; <sup>7</sup>Logistic regression model with standard errors adjusted for clustering by study.

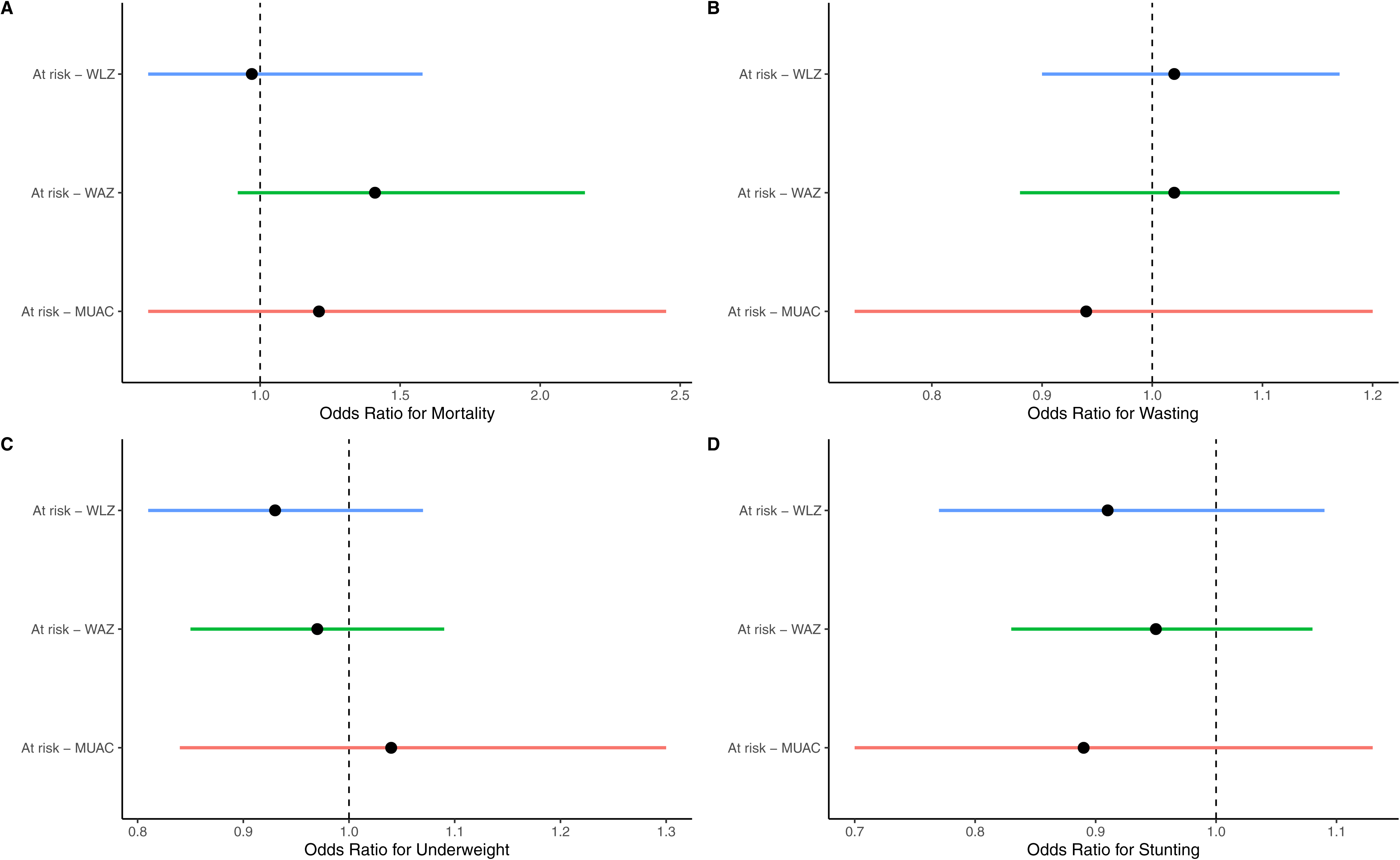

## DISCUSSION

In this pooled analysis including more than 9,000 infants in Burkina Faso at risk of poor growth and development, we found no evidence of an effect of azithromycin for the prevention of mortality or wasting by 6 months of age. In the 2023 revised guidelines, WHO identified this subgroup of infants as a priority population for prevention of wasting.^3^ Children hospitalized with severe acute malnutrition have a large burden of infectious disease.^17^ Children aged 6-59 months who are admitted to outpatient nutritional programs without clinical signs of infection receive routine antibiotics (typically amoxicillin) as part of standard outpatient care.^18^ However, the evidence base is mixed, with some studies showing a benefit of antibiotics for nutritional recovery and prevention of mortality in children with severe acute malnutrition, and others showing no evidence of a difference in outcomes in children receiving amoxicillin compared to placebo.^4,5^ Antibiotics are not routinely used in children aged 6-59 months with moderate acute malnutrition or for prevention of wasting in this age range. Mass distribution of single dose azithromycin has been shown to reduce all-cause mortality in children aged 1-59 months, with some evidence of a stronger effect in infants with underweight and older children with global acute malnutrition.^6,7,19,20^ Limited evidence exists of the role of antibiotics in malnourished infants < 6 months.^21^ Future studies designed specifically to assess the role of antibiotics in this population, and considering other classes of antibiotics, are warranted.

A previous subgroup analysis of the NAITRE trial overall provided limited evidence of an effect of azithromycin in babies with low birthweight or underweight.^12^ In the overall trial populations, there was no evidence of an effect of azithromycin on anthropometric endpoints, including wasting, at 6 months in NAITRE and CHATON.^10,11^ Although the two trials enrolled nearly 55,000 infants, mortality is a rare endpoint, even in high risk subgroups. Here, we pooled data from the two trials to improve statistical power and provide evidence across a wider range of ages of infants under 6 months of age, using the definition of “at risk of poor growth and development” as outlined in the revised WHO guidelines. Although LAZ scores were modestly higher in infants receiving azithromycin compared to placebo, the difference may not be clinically significant and was not apparent when evaluating LAZ dichotomously as stunting. The results of the present analysis suggest that, in line with other subgroups of infants, single dose azithromycin is not effective for preventing mortality or wasting in infants at risk of poor growth and development.

As expected, infants classified as at risk of poor growth and development based on a single measure of WLZ, WAZ, or MUAC had higher risk of mortality, wasting, underweight, and stunting at 6 months of age compared to those not classified at risk of poor growth and development. Low WAZ was the strongest indicator of mortality by 6 months of age and had similar risk of wasting by 6 months as WLZ. Previous work has shown that WAZ and MUAC are strong indicators of risk of mortality in both infants and older children.^22–26^ Inclusion of WAZ as an indicator for identifying children at risk of growth failure and mortality will be critical in screening programs to comprehensively identify those at highest risk of poor outcomes.

There are some limitations to consider in interpretation of this analysis. The NAITRE and CHATON studies were not designed specifically to evaluate antibiotics for infants at risk of poor growth and development. NAITRE had weight-based exclusion criteria that were implemented for safety reasons due to concerns that azithromycin would be more likely to lead to infantile hypertrophic pyloric stenosis in lower-weight infants.^27^ Due to the randomized nature of the trials, this would not be expected to affect internal validity, however it may reduce generalizability. Infants in both trials were recruited via routine health care visits (e.g., following facility birth, vaccination visits) for well-child visits. It is possible that this recruitment strategy selected for lower-risk infants, as those with reduced access to routine healthcare may be more likely to be at risk. Strategies that recruit infants who do not have access to primary care may identify a greater number of infants at risk of poor growth and development. While pooling data from both trials led to a sample size of nearly 10,000 infants at risk of poor growth and development, the mortality probability was approximately 1%, and confidence intervals for mortality analyses were relatively wide. However, wasting was much more common (20% at 6 months), and results were consistent with no evidence of an effect of azithromycin for prevention of wasting in this subgroup of infants. This study provides evidence of the effect of single dose azithromycin on 6-month outcomes. It is possible that other antibiotic regimens, such as amoxicillin which is typically used as part of treatment for children 6-59 months with severe wasting, could have a different effect. Finally, the trials did not follow children beyond 6 months of age, and longer-term effects of early life azithromycin is unknown. However, it is unlikely that a longer-term effect would be observed in the absence of short-term changes.

In this pooled analysis of two randomized placebo-controlled trials that followed nearly identical protocols in Burkina Faso, we did not find evidence to suggest that single dose azithromycin in early infancy prevents mortality or wasting in infants at risk of poor growth and development as defined by WHO. This population of infants has been identified by WHO for evaluation of interventions to reduce further growth deficits and prevent severe wasting, including evaluation of antibiotics. These results do not support implementation of routine azithromycin in infants at risk of poor growth and development.

## Acknowledgements

MB: study design, study implementation, supervision, critical review. MO: study design, study implementation, supervision, critical review. CD: study implementation, supervision, critical review. BC: study design, study implementation, supervision, critical review. TO: study design, study implementation, supervision, critical review. AZ: study design, study implementation, data management, critical review. VB: study design, study implementation, data management, critical review. EL: study design, study implementation, supervision, critical review. HH: study design, study implementation, data management, critical review. BFA: study design, study implementation, data analysis, critical review. TML: study design, study implementation, supervision, critical review. CEO: study design, study implementation, supervision, data analysis, drafting manuscript.

## Data Availability

Data underlying all analyses are publicly available via the trials’ data repositories.

## Funding

The NAITRE and CHATON studies were supported by the Bill and Melinda Gates Foundation (OPP1187628; PI: Lietman). Azithromycin and matching placebo were donated by Pfizer, Inc (New York, NY). The funders played no role in the design, analysis, interpretation, or decision to publish.

## Author Disclosures

None to report.

## Notes

### Competing Interest Statement

The authors have declared no competing interest.

### Clinical Trial

NCT03682654 and NCT03676764

### Author Declarations

The IRB at the University of California San Francisco gave ethical approval for this work. The Comite National d Ethique pour la Recherche en Sante in Ouagadougou Burkina Faso gave ethical approval for this work.

