## Supplemental Tables for "Azithromycin for infants at risk of poor growth and development: a pooled secondary analysis of two randomized controlled trials"

**Supplemental Table 1.** Baseline characteristics by randomized treatment assignment in the NAITRE (age 8-27 days) and CHATON (age 28-83 days) full cohorts

|  | **NAITRE** | | **CHATON** | |
| --- | --- | --- | --- | --- |
|  | Azithromycin | Placebo | Azithromycin | Placebo |
| N | 10,898 | 10,934 | 16,416 | 16,461 |
| Child’s sex, N (%) |  |  |  |  |
| Female | 5,413 (50%) | 5,431 (50%) | 8,045 (49%) | 8,136 (49%) |
| Male | 5,485 (50%) | 5,503 (50%) | 8,371 (51%) | 8,325 (51%) |
| Age in days, median (IQR) | 11 (9 to 15) | 11 (9 to 14) | 46 (35 to 61) | 47 (36 to 62) |
| Season of enrollment |  |  |  |  |
| Rainy (June-October) | 5,217 (48%) | 5,295 (48%) | 7,174 (44%) | 7,279 (44%) |
| Dry (November-May) | 5,681 (52%) | 5,639 (52%) | 9,242 (56%) | 9,182 (56%) |
| Region |  |  |  |  |
| Boucle du Mouhoun | 1,299 (12%) | 1,329 (12%) | 14,231 (87%) | 14,271 (87%) |
| Hauts-Bassins | 5,454 (50%) | 5,465 (50%) | 887 (5%) | 885 (5%) |
| Cascade | 2,009 (18%) | 1,977 (18%) | 1,298 (8%) | 1,305 (8%) |
| Centre | 919 (8%) | 951 (9%) | 0 | 0 |
| Centre Ouest | 1,217 (11%) | 1,211 (11%) | 0 | 0 |
| WLZ, mean (SD) | -0.6 (1.3) | -0.6 (1.3) | -0.2 (1.5) | -0.2 (1.5) |
| WAZ, mean (SD) | -0.6 (0.9) | -0.6 (0.9) | -0.6 (1.2) | -0.6 (1.2) |
| LAZ, mean (SD) | -0.5 (1.0) | -0.5 (1.1) | -0.5 (1.3) | -0.5 (1.3) |
| MUAC, mean (SD) | 10.9 (1.1) | 10.9 (1.1) | 12.1 (1.2) | 12.1 (1.2) |
| At risk of poor growth and development |  |  |  |  |
| Total | 1,928 (18%) | 1,875 (17%) | 2,985 (18%) | 2,940 (18%) |
| By WLZ | 1,475 (14%) | 1,428 (13%) | 1,577 (10%) | 1,542 (9%) |
| By WAZ | 832 (8%) | 785 (7%) | 1,866 (11%) | 1,902 (12%) |
| By MUAC | 0 | 0 | 746 (8%) | 795 (8%) |
